# Vaping and Socioeconomic Inequalities in Smoking Cessation and Relapse: A Longitudinal Analysis of the UK Household Longitudinal Study

**DOI:** 10.1101/2022.08.30.22279385

**Authors:** Iain Hardie, Michael J. Green

**Affiliations:** MRC/CSO Social and Public Health Sciences Unit, University of Glasgow, Glasgow, UK

## Abstract

**Background:** Smoking is a key cause of socioeconomic health inequalities. Vaping is considered less harmful than smoking and has become a popular smoking cessation aid. However, there is currently limited evidence on the impact of vaping on inequalities in smoking.

**Methods:** We used longitudinal data from 25,102 participants in waves 8-10 (2016-2020) of the UK Household Longitudinal Study to examine how vaping affects socioeconomic inequalities in smoking cessation and relapse. Marginal structural models were used to investigate whether vaping mediates or moderates associations between educational attainment and smoking cessation and relapse over time. Multiple Imputation and weights were used to adjust for missing data.

**Results:** Respondents without degrees were less likely to stop smoking than those with a degree (OR: 0.65; 95% CI: 0.54-0.77), and more likely to relapse (OR: 1.67; 95% CI: 1.26-2.23) but regular vaping eliminated the inequality in smoking cessation (OR: 0.99; 95% CI: 0.54-1.82). Sensitivity analyses suggested that this finding did not hold when comparing those with or without any qualifications. Inequalities in smoking relapse did not differ by vaping status.

**Conclusions:** Vaping may help reduce inequalities in smoking cessation between those with and without degree-level education and policy should favour vaping as a smoking cessation aid. Nevertheless, other supports or aids may be needed to reach the most disadvantaged (i.e. those with no qualifications) and to help people avoid relapse after cessation.

**WHAT THIS PAPER ADDS:** *What is already known on this topic?:*

- Socioeconomic inequalities in smoking cessation have narrowed in recent years since e-cigarettes have become more widely available as a cessation aid.
- It is not clear whether this was as a result of increased vaping or other due to other confounding factors.
- Existing research on vaping and socioeconomic inequalities in smoking cessation have been limited to using cross-sectional data.

*What this study adds?:*

- Using longitudinal data, over 2 years of follow-up, our study suggests that increased vaping among those of lower SEP (i.e. without degrees) is likely to have reduced socioeconomic inequalities in smoking cessation.
- However, the positive impact of vaping on inequalities is focused around the upper to middle end of the educational distribution, and does not appear to help the most disadvantaged, or help with inequalities in smoking relapse.

*How this study might affect research, practice or policy:*

- Vaping can most likely have a net positive impact on inequalities in smoking. Policy should favour vaping, although other aids may be needed for the most disadvantaged and to help people avoid smoking relapse.

## INTRODUCTION

Smoking is a leading cause of death and ill-health and contributes substantially to socioeconomic health inequalities in the UK and elsewhere [1-4]. E-cigarettes (i.e. vaping products) offer an alternative method of nicotine delivery to smoking, and are currently the most popular aid for smoking cessation in England, used by around 6% of adults [1]. Vaping is considered to be markedly less harmful than smoking [5, 6] and, whilst more evidence is needed [7-9], current research suggests vaping may be associated with smoking cessation [1, 10-12], and may even be more effective as a cessation aid than nicotine replacement therapy [8]. However, current UK and international evidence also suggests that, among ex-smokers, vaping may increase the risk of a relapse to traditional cigarettes [13, 14], particularly among those vaping infrequently or using less advanced devices [15].

One important aspect of e-cigarette usage relates to its impacts on socioeconomic inequalities. Smoking cessation has tended to be less likely for smokers who are in a more disadvantaged socioeconomic position (SEP), with disadvantaged smokers being less likely to quit and more likely to relapse, but not less likely to want to quit [16-20]. In theory, e-cigarettes may potentially reduce this socioeconomic inequality if they can make smoking cessation more accessible for disadvantaged smokers, but conversely may widen inequalities if vaping exposes people from lower SEP groups to increased risk of relapse to smoking [21, 22]. Analysis of trends in quit success rates suggests that inequalities in smoking cessation have narrowed in recent years since e-cigarettes have become more widely available as a cessation aid [23], but it is not clear whether this was as a result of increased vaping or other due to other confounding factors.

Current evidence on the impact of vaping on socioeconomic inequalities in smoking cessation/relapse is fairly limited. One systematic review suggests that e-cigarette ‘awareness’, ‘ever use’ and ‘current use’ was patterned by a range of sociodemographic factors, but that overall there tended to be a lack of a clear pattern in these outcomes with regards to SEP, particularly in high-quality studies [24]. Data from the Smoking Toolkit Study in England suggests that e-cigarette use increased for all SEP groups from 2014-2019 but was highest among those from lower SEP groups, and there was no differences in post-cessation initiation of e-cigarette use between different SEP groups [22]. Meanwhile, further cross-sectional research using the UK Household Longitudinal Study (UKHLS) suggests that socioeconomic inequalities in smoking cessation (i.e. adult smokers from more advantaged SEP groups being more likely to have quit) were weaker amongst those who vaped [21]. This highlights the potential for e-cigarettes to narrow health inequalities by helping disadvantaged smokers to quit, and suggests that vaping may have contributed to the reduction in inequalities in smoking cessation success rates that has been observed in recent years [23].

The interplay between vaping and smoking can be complex, involving, for example, patterns of dual-use (with or without intentions to quit smoking), switching fully from smoking to vaping, or using vaping as a ‘stepping stone’ to stop smoking and eventually cease nicotine use [1, 9, 25, 26]. However, since smoking is considered to be by far the more harmful behaviour for health [5, 6], inequalities in smoking are of more critical public health importance. With the potential both for inequalities in vaping behaviour and for effects of vaping on cessation and relapse rates it may be helpful to frame the issue in terms of whether vaping mediates or moderates inequalities in smoking cessation and relapse. Thought of this way, it is important to recognise that ‘mediation’ could include ‘suppression’ effects [27], where, for example, vaping might be more common among disadvantaged smokers and might help them quit, thus leading to narrower inequalities in cessation than would be present without access to e-cigarettes. Even without inequalities in vaping, it is possible that vaping could impact inequalities in smoking if it moderates associations between SEP and relapse or cessation [28].

The aim of this study is to assess whether vaping mediates or moderates socioeconomic inequalities in smoking cessation and inequalities in relapse after cessation. Specifically, this study addresses the following questions, over 2 years of follow-up:

Among current smokers:

1. Is SEP associated with vaping?
2. Is vaping associated with smoking cessation?
3. Is SEP associated with smoking cessation?
4. Does vaping mediate or moderate associations between SEP and smoking cessation?

Among ex-smokers:

1. Is SEP associated with vaping?
2. Is vaping associated with relapse to smoking?
3. Is SEP associated with relapse to smoking?
4. Does vaping mediate or moderate associations between SEP and relapse to smoking?

## METHODS

### Data and Sample

Analyses used longitudinal data from annual interviews in waves 8-10 of the UK Household Longitudinal Study (UKHLS), a nationally representative household panel study, which is based on a clustered-stratified probability sample of around 40,000 UK households [29]. It began its first wave of data collection in 2009-2011, and individuals from the same households are visited each year for annual interviews, conducted either face-to-face or online. Our analysis primarily used data from wave 8 (collected 2016-2018), wave 9 (collected 2017-2019) and wave 10 (collected 2018-2020), although some information from earlier waves was used where applicable (see below). Waves 8-10 were selected as they included more detailed categorisations of vaping status than previous waves, and are the most recent waves which were unaffected by the COVID-19 pandemic.

Using smoking status at wave 8 as a baseline, smoking cessation and relapse were then measured over the following two years (waves 9-10). UKHLS respondents were included in our analysis if they met the following inclusion criteria: (1) were interviewed at wave 8, (2) had a valid, non-missing wave 8 weight, and (3) had data on smoking status at waves 9 or 10. This gave a final primary sample of 25,102 individual respondents (see Supporting Information Appendix A for details of sample size, exclusions and missing data). All analyses were conducted using Stata/MP 17.0. Weights were applied to adjust for survey design and survey non-response. Item non-response was dealt with via multiple imputation, using chained equations [30], with 50 imputations added (see Supporting Information Appendix A for details of missingness across variables).

### Measured Variables

Our sample was stratified by baseline smoking status (1=never smoker, 2=ex-smokers, 3=current smoker; defined using self-reports at wave 8 and earlier waves). Ex-smokers were those who did not smoke at wave 8 but had previously smoked regularly (i.e. at least once per day). Outcomes were binary indicators measuring: (1) smoking cessation by wave 9 or 10 among wave 8 current smokers (0=no, 1=yes), and (2) smoking relapse by wave 9 or 10 among wave 8 ex-smokers (0=no, 1=yes). Our main exposure variable, SEP, was represented using educational attainment (0=degree, 1=no degree). Wave 8 self-reported regular (i.e. at least weekly) vaping status was defined as a mediator (0=not regular vaper, 1=regular vaper).

Causal relationships between SEP, vaping and smoking cessation/relapse are complex, with various potential confounders at different stages of the causal pathway (see Figure 1). Consequently, our analysis included: (1) a list of exposure-outcome (and exposure-mediator) confounders, i.e. potential determinants of both exposure (wave 8 SEP), mediator (wave 8 vaping) and outcome (smoking cessation/relapse at waves 9 or 10); and (2) a list of mediator-outcome confounders, i.e. potential determinants of both mediator (wave 8 vaping) and outcome (smoking cessation/relapse at waves 9 or 10), some of which may have been caused by the exposure (wave 8 SEP). The exposure-outcome variables included in our analysis were as follows: sex (0=male, 1=female), age group (1=16-24, 2=25-34, 3=35-44, 4=45-54, 5=55+), UK country (1=England, 2=Wales, 3=Scotland, 4=Northern Ireland), ethnicity (0=white, 1=non-white), and rurality (0=rural, 1=urban). The mediator outcome variables were as follows: partner status (0=in couple, 1=single), has kids (0= no, 1=yes) housing tenure (0=owner, 1=renter), National Statistics Socio-economic Classification (NSSEC) (1=management/professional, 2=Intermediate, 3=Routine, 4=Not in paid employment), has long-standing illness (0=no, 1=yes), vaping history (0=does not vape at all at wave 7, 1=vapes at all at wave 7) mental health (measured by General Health Questionnaire [GHQ]) (1=GHQ <4, 2=GHQ 4+), poverty status (0=not in poverty, 1=in poverty), age started smoking (0=0-15, 1=16-16, 2=19-25, 3=>25), and smoking history - mean number of cigarettes per day across waves or when last smoked regularly (0 = 0-10, 1 = 11-20, 2 = >20). With the exception of the vaping and smoking history variables, all exposure-outcome variables and mediator-outcome variables were measured at wave 8, though values from wave 7 were used if wave 8 data was missing.

**Figure 1.**
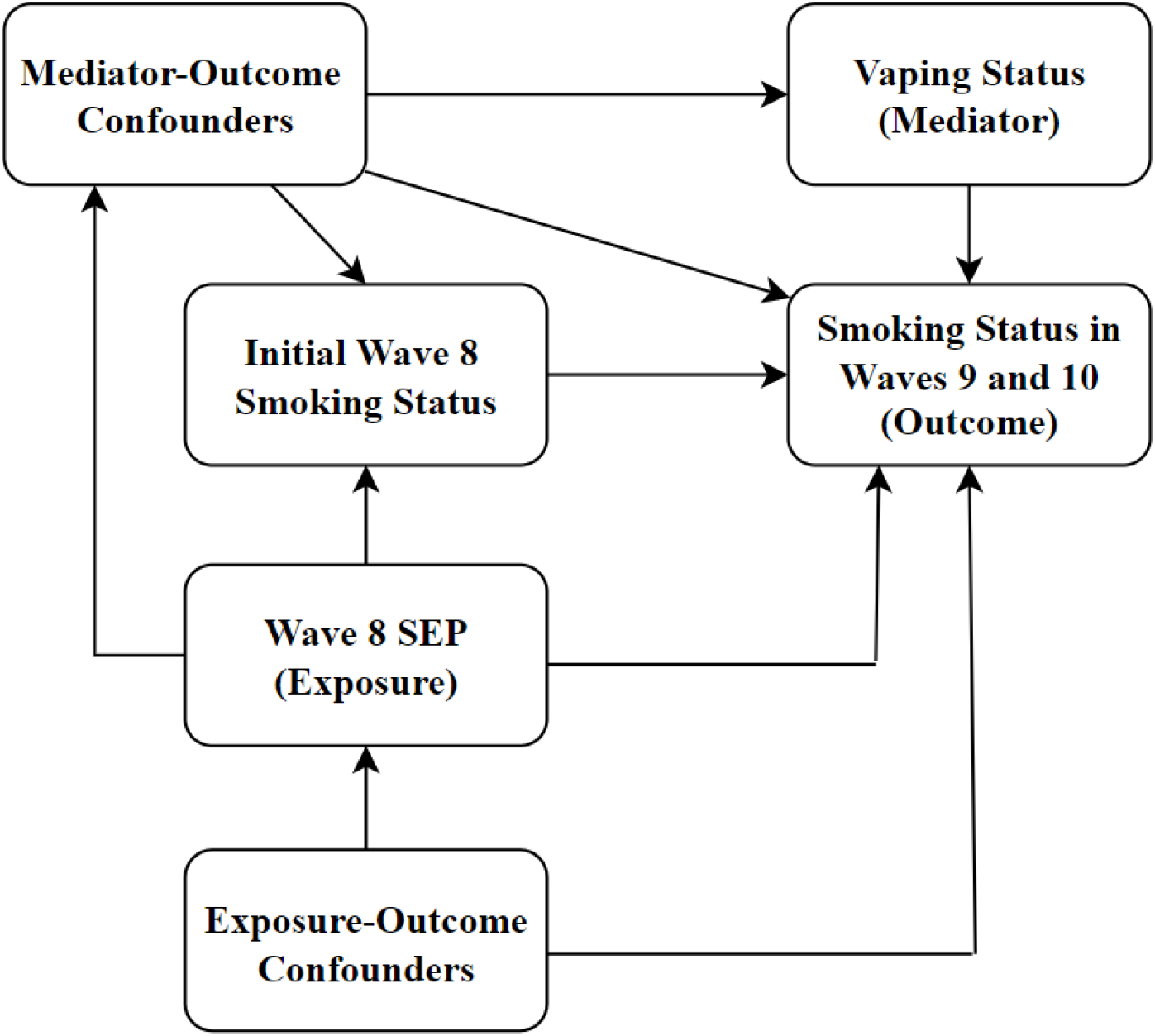
Causal Diagram of the Relationship between SEP, Vaping and Smoking Cessation/Relapse

### Statistical Analysis

Our analysis plan was preregistered using Open Science Framework (available here: https://osf.io/e3z8q), and our reporting is consistent with STROBE guidelines (see Supporting Information Appendix B). First, we used logistic regression to estimate unadjusted associations between the variables of interest in each of our four research questions. These unadjusted associations may be subject to collider bias [31, 32], because the data is stratified by wave 8 smoking status, which is potentially determined by both: (1) the exposure variable, and (2) other variables determining cessation/relapse. This is shown in Figure 1 – wave 8 smoking status is determined by both SEP and mediator-outcome confounders.

Second, to account for this, we used inverse-probability weighted marginal structural models to estimate controlled direct effects (CDE) of SEP on smoking cessation/relapse, controlling for observed confounding [33]. The CDE represents the effect of the exposure, with mediators set to a particular level (e.g. setting wave 8 status to either current or ex-smoking, and to either regular vaping or not regular vaping). Weights were calculated within each imputed data set and final results were aggregated across imputed datasets using Rubin’s rules [30]. These models aim to remove any imbalance of observed confounders across exposure levels that is not caused by the exposure. CDE estimates account for interactions between the exposure and the mediators and may therefore vary depending on the values mediators are set to [33]. As explained below, some of our CDE estimates treat wave 8 smoking status as the only mediator, so provide estimates with wave 8 smoking set to either current or ex-smoking (to get separate estimates for cessation and relapse). Later estimates include vaping as an additional mediator and compare estimates with vaping set to regular or not regular vaping. We estimate effects across two waves of follow-up using a discrete-time, event history approach, with up to two rows of data for waves 9 and 10; the wave 10 row is censored if cessation/relapse occurs at wave 9. Thus, odds ratios (ORs) can be interpreted as the hazard or risk of cessation/relapse in a given year, if this has not already occurred.

For research question 1 (Is SEP associated with vaping?), we created a weight to estimate the CDE of education with wave 8 smoking status set to either current or ex-smoking. This adjusts for (exposure-outcome) confounders of education, vaping and smoking through follow up, and for (mediator-outcome) confounders of smoking status at wave 8, vaping and smoking through follow-up. A similar set of weights were then used for research question 2 (Is vaping associated with smoking cessation?), but with vaping treated as the exposure rather than education, and cessation/relapse as the outcome. For research question 3 (Is SEP associated with smoking cessation/relapse?) the same weights as research question 1 were used to estimate the CDE of education on smoking cessation/relapse. Finally, for research question 4 (does vaping mediate or moderate the associations between SEP and smoking cessation/relapse?), the same inverse probability weights used for research questions 1 and 2 were used, but with an additional step of weighting to account for regular vaping as the mediator. We produced separate CDE estimates for effects of education on cessation/relapse with vaping status set to either regular or not regular vaping. For full details of the process of creating the weights and running the modelling for each research question see Supporting Information Appendix C.

Additional sensitivity analyses were performed as specified in our preregistered plan. Each tests the specificity of our results by repeating analyses using alternative categorisations or measures. Firstly, vaping status was recoded to indicate any vaping (0=non-vaper, 1=infrequent or regular vaper). Next we used two binary classifications based on the UK National Statistics Socioeconomic Classification (NSSEC) for occupations as our main SEP measure (0=management/professional, 1=not management/professional; and 0=in paid employment, 1=not in paid employment), with education re-classified as an exposure-outcome confounder. This assesses whether there is evidence for any additional effect of a more proximal SEP measure, over and above the effect of the education measure used in the main analyses. Finally, analyses were repeated with education recoded to indicate possession of any qualifications (0=has qualifications, 1=no qualifications).

## RESULTS

### Descriptive Statistics

Descriptive statistics showing sociodemographic patterning of our sample by wave 8 smoking and vaping status are provided in Table 1. Of the total sample, 16.1% were smokers and 30.1% ex-smokers at wave 8. Smoking was disproportionately prevalent among people who had no degree, as well as among those who were single, renting, younger, in urban areas, in poverty, with a longstanding illness or who had higher GHQ scores. Regular vaping was rare, with 4.0% of the sample vaping at least weekly. However, vaping was more prevalent amongst those who had ever smoked, with 8.4% of ex-smokers and 8.8% of current smokers being regular vapers compared to 0.2% of never smokers. Vaping was also disproportionately prevalent among those with no degree and those who were male, aged 25-34, white, renting, in urban areas or with kids in their household.

**Table 1.** Sociodemographic Patterning of Sample by Wave 8 Smoking and Vaping Status

| Covariates | Unweighted N (Weighted %) |  |  |  |  |  |
| --- | --- | --- | --- | --- | --- | --- |
|  | Total | Never Smokers | Ex-Smokers | Current Smokers | Non-Reg Vapers | Reg Vapers |
|  | 25,102 (100%) | 13,511 (53.7%) | 7,724 (30.1%) | 3,867 (16.1%) |  |  |
| **Vaping** |  |  |  |  |  |  |
| Not Reg Vaper | 24,136 (96.0%) | 13,489 (99.8%) | 7,134 (91.6%) | 3,513 (91.2%) |  |  |
| Reg Vaper | 966 (4.0%) | 22 (0.2%) | 590 (8.4%) | 354 (8.8%) |  |  |
| **Degree** |  |  |  |  |  |  |
| No Degree | 15,743 (63.8%) | 7,603 (57.3%) | 5,090 (66.6%) | 3,049 (80.4%) | 15,036 (63.3%) | 706 (75.0%) |
| Has Degree | 9,359 (36.2%) | 5,907 (42.7%) | 2,634 (33.4%) | 818 (19.6%) | 9,100 (36.7%) | 259 (25.0%) |
| **Sex** |  |  |  |  |  |  |
| Male | 11,035 (47.8%) | 5,394 (45.3%) | 3,800 (51.1%) | 1,841 (49.7%) | 10,544 (47.5%) | 491 (54.1%) |
| Female | 14,067 (52.2%) | 8,117 (54.7%) | 3,924 (48.9%) | 2,026 (50.3%) | 13,592 (52.5%) | 475 (45.9%) |
| **Age** |  |  |  |  |  |  |
| 16-24 | 1,601 (12.8%) | 1,062 (16.4%) | 153 (4.2%) | 386 (17.1%) | 1,543 (13.0%) | 58 (9.7%) |
| 25-34 | 2,682 (13.1%) | 1,520 (13.6%) | 561 (9.3%) | 601 (18.4%) | 2,524 (12.7%) | 158 (20.9%) |
| 35-44 | 4,028 (14.9%) | 2,235 (14.8%) | 1,083 (14.3%) | 710 (16.5%) | 3,821 (14.7%) | 207 (19.6%) |
| 45-54 | 5,037 (17.9%) | 2,780 (17.7%) | 1,427 (17.7%) | 830 (18.7%) | 4,804 (17.7%) | 233 (21.9%) |
| 55+ | 11,754 (41.3%) | 5,914 (37.5%) | 4,500 (54.6%) | 1,340 (29.3%) | 11,444 (41.9%) | 310 (27.9%) |
| **Ethnicity** |  |  |  |  |  |  |
| White | 22,087 (92.0%) | 11,468 (89.3%) | 7,230 (95.6%) | 3,389 (94.2%) | 21,199 (91.9%) | 888 (95.3%) |
| Non-White | 3,015 (8.0%) | 2,043 (10.7%) | 494 (4.4%) | 478 (5.8%) | 2,937 (8.1%) | 78 (4.7%) |
| **NSSEC** |  |  |  |  |  |  |
| Management/Professional | 6,426 (24.7%) | 4,066 (29.4%) | 1,748 (22.2%) | 612 (13.8%) | 6,207 (24.8%) | 219 (21.5%) |
| Intermediate | 3,516 (13.7%) | 2,016 (15.0%) | 985 (12.1%) | 515 (12.1%) | 3,348 (13.6%) | 168 (15.8%) |
| Routine | 4,569 (20.3%) | 2,206 (18.6%) | 1,257 (17.9%) | 1,106 (30.4%) | 4,313 (20.0%) | 256 (28.6%) |
| Not in paid Employment | 10,591 (41.3%) | 5,223 (36.9%) | 3,734 (47.8%) | 1,634 (43.7%) | 10,268 (41.6%) | 323 (34.0%) |
| **Single in Household** |  |  |  |  |  |  |
| No | 16,834 (59.7%) | 9,248 (59.1%) | 5,499 (67.5%) | 2,087 (46.8%) | 16,218 (59.7%) | 616 (57.8%) |
| Yes | 8,268 (40.3%) | 4,263 (40.9%) | 2,225 (32.5%) | 1,780 (53.2%) | 7,918 (40.3%) | 350 (42.2%) |
| **Kids in Household** |  |  |  |  |  |  |
| No | 18,681 (76.2%) | 9,899 (76.8%) | 5,975 (76.7%) | 2,807 (72.9%) | 18,012 (76.4%) | 669 (69.8%) |
| Yes | 6,421 (23.8%) | 3,612 (23.2%) | 1,749 (23.3%) | 1,060 (27.1%) | 6,124 (23.6%) | 297 (30.2%) |
| **Tenure** |  |  |  |  |  |  |
| Owner | 18,960 (67.9%) | 11,057 (75.0%) | 5,940 (69.4%) | 1,963 (41.5%) | 18,381 (68.7%) | 579 (49.1%) |
| Renter | 6,142 (32.1%) | 2,454 (25.0%) | 1,784 (30.6%) | 1,904 (58.5%) | 5,755 (31.3%) | 387 (50.9%) |
| **Rural/Urban** |  |  |  |  |  |  |
| Rural | 6,678 (24.2%) | 3,624 (24.4%) | 2,224 (25.8%) | 830 (20.5%) | 6,477 (24.4%) | 201 (19.8%) |
| Urban | 18,424 (75.8%) | 9,887 (75.6%) | 5,500 (74.2%) | 3,037 (79.5%) | 17,659 (75.6%) | 765 (80.2%) |
| **Has Longstanding Illness** |  |  |  |  |  |  |
| No | 15,596 (63.1%) | 9,028 (68.3%) | 4,293 (55.5%) | 2,275 (59.8%) | 15,013 (63.3%) | 583 (59.4%) |
| Yes | 9,506 (36.9%) | 4,483 (31.7%) | 3,432 (44.5%) | 1,592 (40.2%) | 9,123 (36.7%) | 383 (40.6%) |
| **In Poverty** |  |  |  |  |  |  |
| No | 21,951 (87.2%) | 11,953 (88.7%) | 6,845 (88.0%) | 3,153 (80.6%) | 21,105 (87.3%) | 846 (85.7%) |
| Yes | 3,151 (12.8%) | 1,558 (11.3%) | 879 (12.0%) | 714 (19.4%) | 3,031 (12.7%) | 120 (14.3%) |
| **GHQ** |  |  |  |  |  |  |
| <4 (less distressed) | 20,519 (80.7%) | 11,278 (83.0%) | 6,356 (80.8%) | 2,885 (73.1%) | 19,759 (80.9%) | 760 (77.4%) |
| 4+ (more distressed) | 4,583 (19.3%) | 2,233 (17.0%) | 1,368 (19.2%) | 981 (26.9%) | 4,377 (19.1%) | 206 (22.6%) |
| **Wave 7 E-Cigs Ever Use** |  |  |  |  |  |  |
| No | 23,331 (92.6%) | 13,423 (99.3%) | 7,085 (91.0%) | 2,823 (73.1%) | 23,033 (95.2%) | 298 (31.4%) |
| Yes | 1,771 (7.4%) | 88 (0.7%) | 639 (9.0%) | 1,044 (26.9%) | 1,103 (4.8%) | 668 (68.6%) |
| **Current/ex-smoker mean #**<br>**cigs per day across waves** |  |  |  |  |  |  |
| <11 | 5,535 (47.9%) |  | 3,393 (44.2%) | 2,142 (54.8%) | 5,130 (48.5%) | 405 (41.7%) |
| 11-20 | 4,539 (39.2%) |  | 3,074 (39.7%) | 1,465 (38.2%) | 4,142 (38.8%) | 398 (43.3%) |
| >20 | 1,517 (12.9%) |  | 1,257 (16.1%) | 260 (7.0%) | 1,376 (12.7%) | 141 (15.0%) |
| **Current/ex-smoker age**<br>**started smoking** |  |  |  |  |  |  |
| Aged <16 | 4,552 (41.6%) |  | 2,782 (37.3%) | 1,770 (49.6%) | 4,131 (41.1%) | 420 (46.6%) |
| Aged 16-19 | 5,010 (42.7%) |  | 3,495 (45.0%) | 1,515 (38.4%) | 4,627 (43.0%) | 383 (39.4%) |
| Aged 19-25 | 1,568 (12.3%) |  | 1,127 (13.8%) | 441 (9.4%) | 1,455 (12.3%) | 113 (11.5%) |
| Aged >25 | 461 (3.5%) |  | 320 (3.9%) | 141 (2.7%) | 433 (3.6%) | 28 (2.6%) |

Notes: Multiple imputed data with 50 imputations added. Weighting uses UKHLS wave 8 sampling weight.

### Effects of SEP on Regular Vaping

Table 2 shows estimated effects of education on vaping among wave 8 current smokers and ex-smokers. Both unadjusted and adjusted CDE estimates are provided. Among current smokers, having no degree was also associated with regular vaping, but confidence intervals over-lapped the null in both the unadjusted (OR: 1.28; 95% CI: 0.93-1.76) and adjusted (OR: 1.24; 95% CI: 0.87-1.78) models. Among ex-smokers, having no degree was associated with increased odds of regular vaping in both unadjusted (OR: 1.27; 95% CI: 1.02-1.60) and adjusted (OR: 1.66; 95% CI: 1.33-2.07) models.

**Table 2.** Estimated Effects of SEP on Regular Vaping Among Current Smokers and Ex-Smokers

|  | **Unadjusted Association**<br>**between Having No Degree and**<br>**Regular Vaping** | **Controlled Direct Effect of**<br>**Having No Degree on Regular**<br>**Vaping** |
| --- | --- | --- |
|  | OR [95% CI] | OR [95% CI] |
| **Wave 8 Regular Vaping (Current Smokers)**<br>[Reference: Degree]<br>No Degree | 1.28 [0.93-1.76] | 1.24 [0.87-1.78] |
| **Wave 8 Regular Vaping (Ex-Smokers)**<br>[Reference: Degree]<br>No Degree | 1.27 [1.02-1.60] | 1.66 [1.33-2.07] |

Notes: OR = odds ratio and 95% CI = 95% confidence interval. Regular vaping is defined as vaping at least weekly. The unadjusted association uses UKHLS wave 8 sampling weights to account for survey design and non-response but does not adjust for any confounders. The controlled direct effect uses inverse probability weighted marginal structural modelling to additionally adjust for exposure-outcome confounders and mediator-outcome confounders.

### Effects of Regular Vaping on Smoking Cessation/Relapse

Table 3 shows estimated effects of regular vaping on smoking cessation/relapse, again providing both unadjusted and adjusted CDE estimates. Regular vaping was associated with increased odds of smoking cessation among wave 8 current smokers (OR: 1.28; 95% CI: 1.03-1.59) but this was attenuated after adjusting for observed confounding (OR: 1.13; 95% CI: 0.82-1.55). Among wave 8 ex-smokers, regular vaping was associated with increased odds of smoking relapse in both unadjusted (OR: 2.75; 95% CI: 2.02, 3.73) and adjusted (OR: 2.97; 95% CI: 2.10-4.22) models.

**Table 3.**
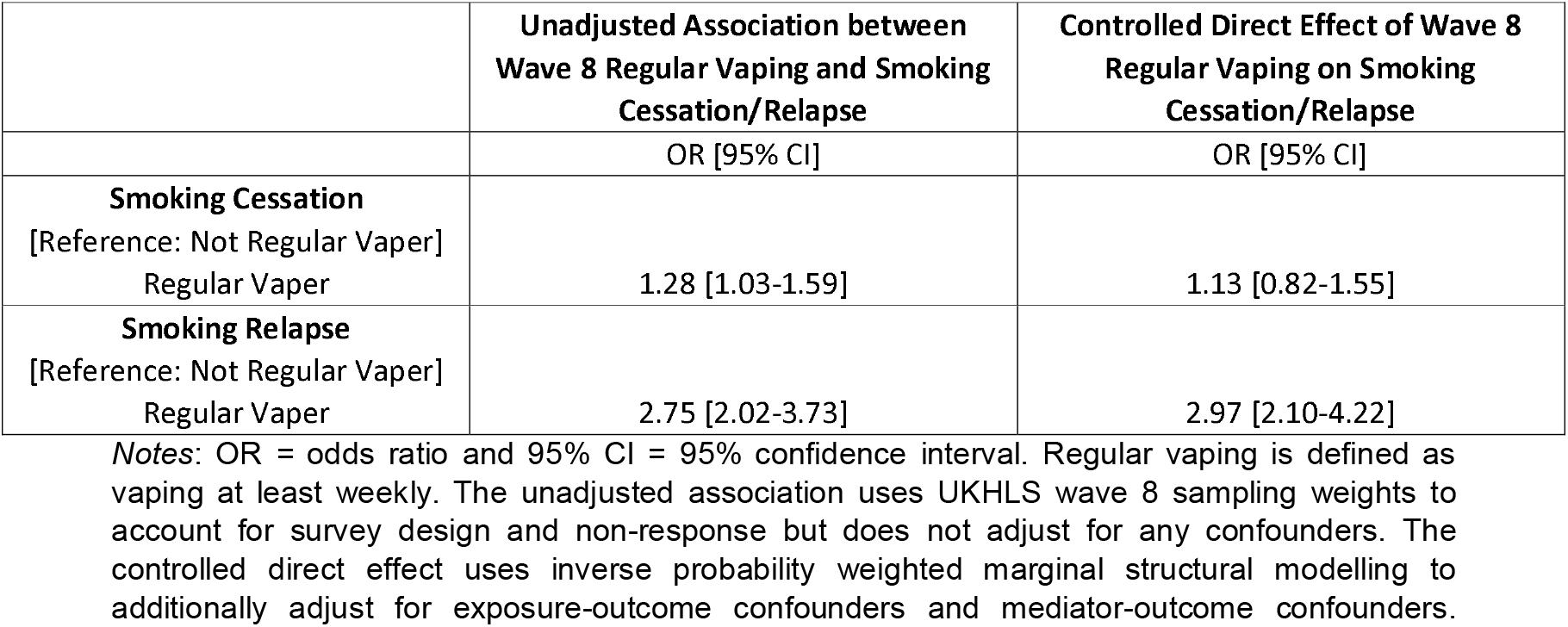
Estimated Effects of Regular Vaping on Smoking Cessation/Relapse

|  | **Unadjusted Association between**<br>**Wave 8 Regular Vaping and Smoking**<br>**Cessation/Relapse** | **Controlled Direct Effect of Wave 8**<br>**Regular Vaping on Smoking**<br>**Cessation/Relapse** |
| --- | --- | --- |
|  | OR [95% CI] | OR [95% CI] |
| **Smoking Cessation**<br>[Reference: Not Regular Vaper]<br>Regular Vaper | 1.28 [1.03-1.59] | 1.13 [0.82-1.55] |
| **Smoking Relapse**<br>[Reference: Not Regular Vaper]<br>Regular Vaper | 2.75 [2.02-3.73] | 2.97 [2.10-4.22] |

Notes: OR = odds ratio and 95% CI = 95% confidence interval. Regular vaping is defined as vaping at least weekly. The unadjusted association uses UKHLS wave 8 sampling weights to account for survey design and non-response but does not adjust for any confounders. The controlled direct effect uses inverse probability weighted marginal structural modelling to additionally adjust for exposure-outcome confounders and mediator-outcome confounders.

### Effects of SEP, and its Interaction with Regular Vaping, on Smoking Cessation/Relapse

Table 4 shows the relationship between SEP and smoking cessation and relapse, with unadjusted associations, CDE effect estimates adjusting for confounding but not for vaping, and CDE estimates dependent on regular vaping status. Among wave 8 current smokers, having no degree was associated with reduced odds of smoking cessation. This was consistent across both the unadjusted (OR: 0.62; 95% CI: 0.52-0.73) and the confounder-adjusted model (OR: 0.65; 95% CI: 0.54-0.77). A similar relationship was present among those who were not regular vapers (OR: 0.62; 95% CI: 0.50-0.76), but the association disappeared for regular vapers (OR: 0.99; 95% CI: 0.54-1.82).

**Table 4.** Estimated Effects of SEP on Smoking Cessation/Relapse, With and Without Interaction by Regular Vaping Status

|  | **Unadjusted Association**<br>**between Having No Degree**<br>**and Smoking**<br>**Cessation/Relapse** | **Controlled Direct Effect of**<br>**Having No Degree on**<br>**Smoking Cessation/Relapse** | **Controlled Direct Effect of**<br>**Having No Degree on**<br>**Smoking Cessation/Relapse**<br>**Among Non-Regular Vapers** | **Controlled Direct Effect of**<br>**Having No Degree on**<br>**Smoking Cessation/Relapse**<br>**Among Regular Vapers** |
| --- | --- | --- | --- | --- |
|  | OR [95% CI] | OR [95% CI] | OR [95% CI] | OR [95% CI] |
| **Smoking Cessation**<br>[Reference: Degree]<br>No Degree | 0.62 [0.52-0.73] | 0.65 [0.54-0.77] | 0.62 [0.50-0.76] | 0.99 [0.54-1.82] |
| **Smoking Relapse**<br>[Reference: Degree]<br>No Degree | 1.28 [0.93-1.75] | 1.67 [1.26-2.23] | 1.51 [1.00-2.28] | 1.83 [0.82-4.08] |

Notes: OR = odds ratio and 95% CI = 95% confidence interval. Regular vaping is defined as vaping at least weekly. The unadjusted association uses UKHLS wave 8 sampling weights to account for survey design and non-response but does not adjust for any confounders. The controlled direct effect uses inverse probability weighted marginal structural modelling to additionally adjust for exposure-outcome confounders and mediator-outcome confounders.

Among wave 8 ex-smokers, having no degree was associated with raised risk of smoking relapse, and this was more strong in the confounder-adjusted model (OR: 1.67; 95% CI: 1.26-2.23), than in the unadjusted model (OR=1.28; 95% CI: 0.93-1.75). After regular vaping was included, the association remained present among regular vapers (OR: 1.83; 95% CI: 0.82-4.08) and those who were not regular vapers (OR: 1.51; 95% CI: 1.00-2.28).

### Sensitivity Analysis

Findings from sensitivity analyses in which vaping status was recoded to include infrequent vapers and regular vapers were consistent with the main analysis (see Supporting Information Appendix D). Analyses using occupational class (NSSEC) suggested little remaining socioeconomic inequality in cessation/relapse after adjusting for educational attainment (see Supporting Information Appendices E and F). Nevertheless, despite wide confidence intervals, both analyses showed cessation as being less likely in disadvantaged occupations, with a similar association for those who did not regularly vape, while for regular vapers the association had reversed in direction. One other difference worth noting, is that respondents who were not in employment had lower odds of vaping among both current and ex-smokers than those who were in employment.

Recoding our education measure to indicate no qualifications produced notably different findings (see Supporting Information Appendix G). Respondents with no qualifications were less likely to be regular vapers (unadjusted OR: 0.69; 95% CI: 0.54-0.87; OR: 0.86; 95% CI: 0.67-1.11) than those with qualifications. Moreover, while smoking cessation was less likely among those with no qualifications this association was present among regular vapers (OR: 0.30; 95% CI: 0.14-0.65) and those who were not regular vapers (OR: 0.75; 95% CI: 0.56-0.99). Together with our main analyses, this suggests a non-linear relationship, whereby vaping may help reduce socioeconomic inequalities in smoking cessation at the middle to upper end of the educational distribution (i.e. between those with and without degrees), but is unlikely to help reduce inequalities at the lower end of the educational distribution (i.e. between those with and without any qualifications).

## DISCUSSION

This study has examined the impact of vaping on socioeconomic inequalities in smoking cessation and relapse using annual data from a large and representative UK survey, the UK Household Longitudinal Survey (UKHLS), spanning from 2016 to early 2020. Our findings suggest that smokers with lower educational attainment were less likely to stop smoking, but this inequality was not present among smokers who vaped regularly. However, vaping only appeared to alleviate inequalities when comparing those at the top of the educational distribution (those with degrees) to those in the middle or bottom (those without degrees). It did not appear to alleviate inequalities at the lower end of the distribution, between those with no educational qualifications and those who did have some. With regards to smoking relapse, our findings suggest that ex-smokers with less education were more likely to relapse but this inequality was present regardless of whether ex-smokers vaped regularly or not.

Importantly, if e-cigarettes can be particularly useful in helping people from lower SEP groups to quit smoking, then this could lead to long-term reductions in health inequalities. Overall, literature on quit success rates in England suggests that socioeconomic inequalities in cessation have been narrowing in recent years [23]. Our findings suggest that increased vaping among those of lower SEP (i.e. without degrees) is likely to have contributed positively to this. We confirm previous cross-sectional research where inequalities were found to be weaker among adult vapers [21], but our study extends this finding with longitudinal data, and demonstrates that the impact of vaping on inequalities is focused around the upper to middle end of the educational distribution, but does little to help those who are most disadvantaged, or to address inequalities in relapse among ex-smokers.

Our study does have some limitations. Firstly, whilst we adjust for many relevant confounders, causal interpretation is based on assumptions of no unmeasured confounding. Given that our analysis was stratified by wave 8 smoking status, this includes unmeasured confounding of smoking at UKHLS wave 8 and through follow up in waves 9 and 10 (i.e. any unmeasured determinant of continued smoking). One obvious candidate for an unmeasured confounder is residual differences in smoking history, which we did adjust for, but the measures were crude (being based on limited data from earlier surveys) and may not fully reflect differences in smoking history between smoking/vaping categories. It is plausible that bias arising from this, for example, may have contributed to the observed association between vaping and greater risk of smoking relapse. An additional limitation is the available data did not distinguish between different types of vaping devices or different motivations for e-cigarette use (e.g. price, quitting smoking, convenience etc.).

Despite these limitations, our findings have some important implications. Whilst inequalities in smoking cessation have previously been a very intractable problem, the results of this study suggest that vaping may help alleviate inequalities between those with and without degrees. This suggests that regulations and policy relating to e-cigarette use should favour and enable vaping as a tool for smoking cessation, especially as vaping may not only help alleviate socioeconomic inequalities, but may also aid cessation more than other tools like nicotine replacement therapy [8]. There are concerns about a potential ‘gateway effect’ between vaping and smoking uptake but latest evidence suggest this is unlikely [34]. Our findings did not show that vaping helped with inequalities between those with and without any qualifications, or with inequalities in smoking relapse, so other types of smoking cessation aids or support may be of more use to those who are most disadvantaged (i.e. with no qualifications), and may be needed to help maintain cessation. Nonetheless, a reduction in inequalities in smoking cessation is significant and likely means that vaping can have a net positive impact on inequalities in smoking.

## Data Availability

The dataset used for this analysis, the UK Household Longitudinal Study, aka Understanding Society, is an initiative funded by the Economic and Social Research Council and various Government Departments, with scientific leadership by the Institute for Social and Economic Research, University of Essex, and survey delivery by NatCen Social Research and Kantar Public. The research data are distributed by the UK Data Service (SN 6614).

https://beta.ukdataservice.ac.uk/datacatalogue/series/series?id=2000053#!/abstract

## SUPPORTING INFORMATION APPENDIX A

**Figure S1.**
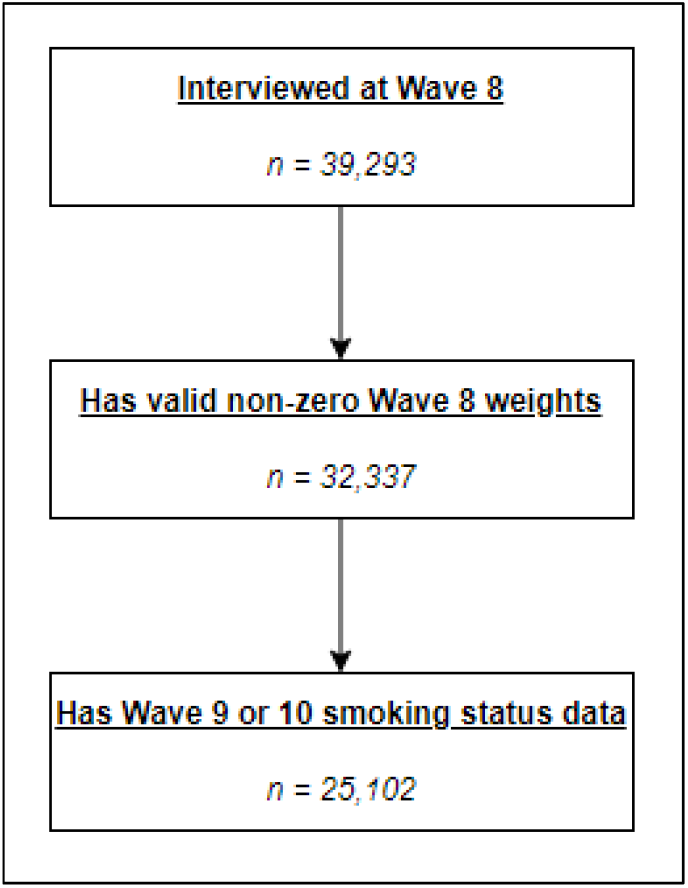
Flowchart showing sample size and missing data at each stage of inclusion criteria

**Table S1.**
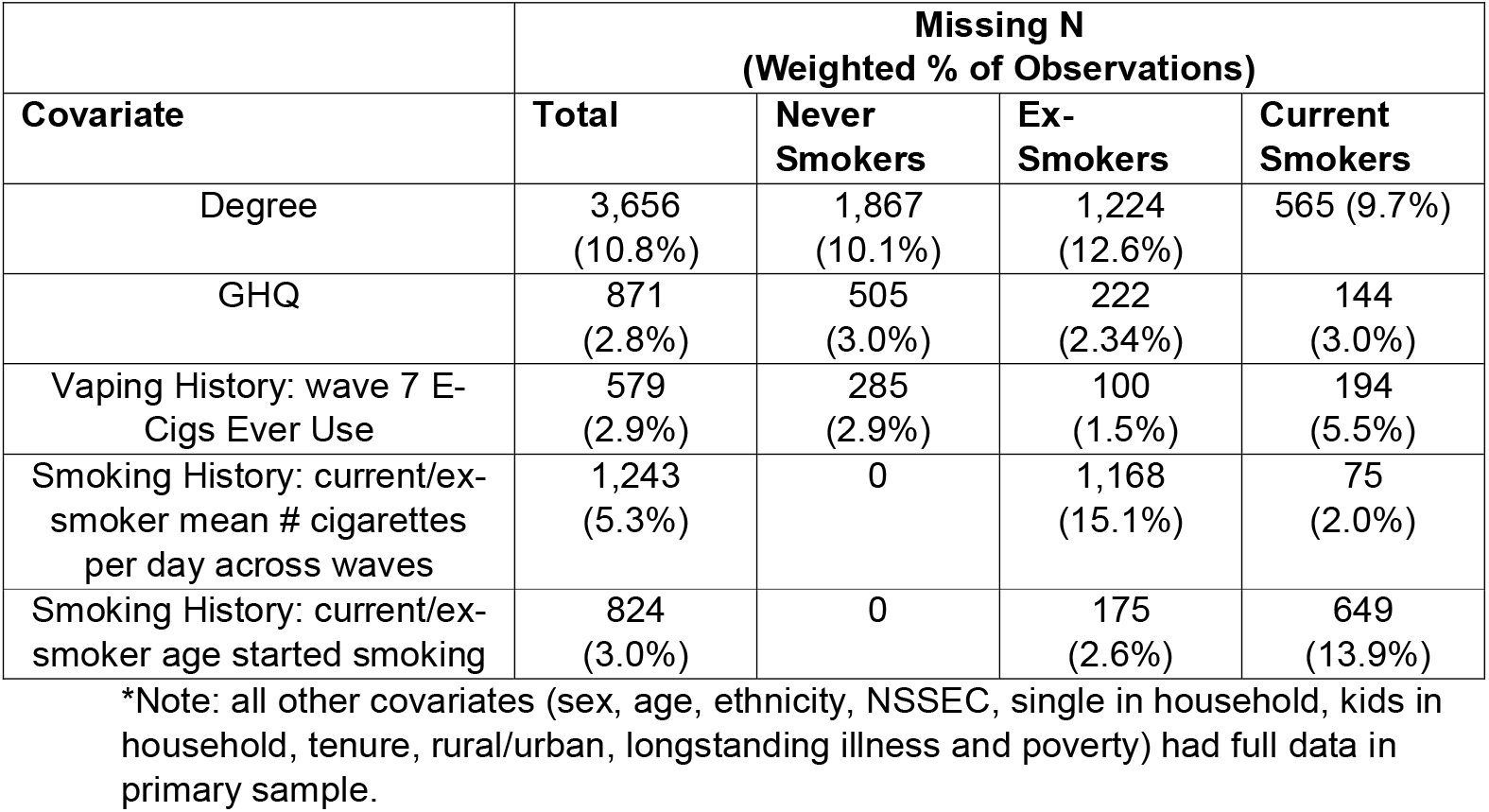
Missingness Across Covariates in Final Primary Sample

| Covariate | Missing N<br>(Weighted % of Observations) |  |  |  |
| --- | --- | --- | --- | --- |
|  | Total | Never Smokers | Ex-Smokers | Current Smokers |
| Degree | 3,656<br>(10.8%) | 1,867<br>(10.1%) | 1,224<br>(12.6%) | 565 (9.7%) |
| GHQ | 871<br>(2.8%) | 505<br>(3.0%) | 222<br>(2.34%) | 144<br>(3.0%) |
| Vaping History: wave 7 E-Cigs Ever Use | 579<br>(2.9%) | 285<br>(2.9%) | 100<br>(1.5%) | 194<br>(5.5%) |
| Smoking History: current/ex-smoker mean # cigarettes per day across waves | 1,243<br>(5.3%) | 0 | 1,168<br>(15.1%) | 75<br>(2.0%) |
| Smoking History: current/ex-smoker age started smoking | 824<br>(3.0%) | 0 | 175<br>(2.6%) | 649<br>(13.9%) |

\*Note: all other covariates (sex, age, ethnicity, NSSEC, single in household, kids in household, tenure, rural/urban, longstanding illness and poverty) had full data in primary sample.

## SUPPORTING INFORMATION APPENDIX B

STROBE Checklist of items that should be included in reports of observational studies

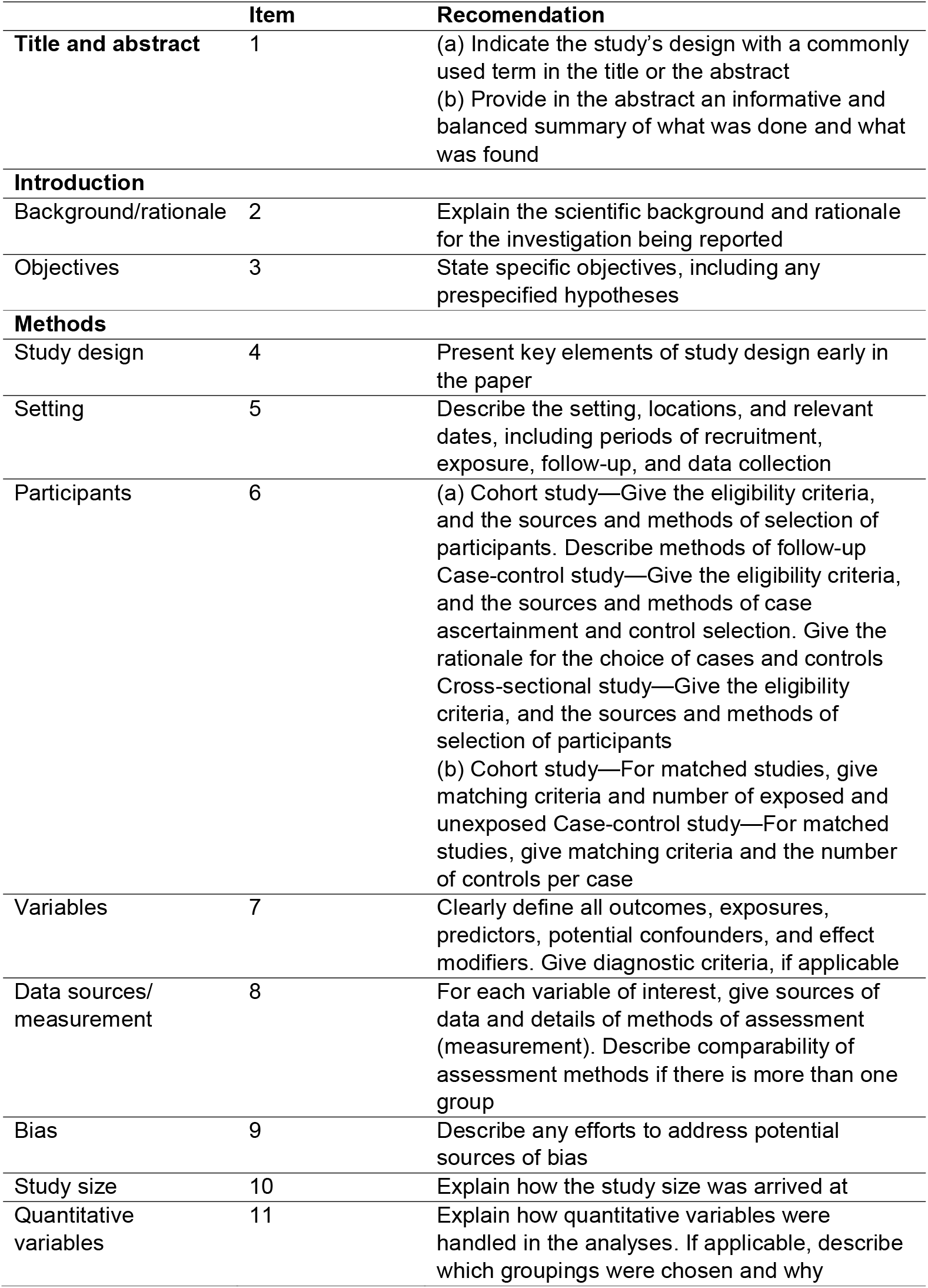
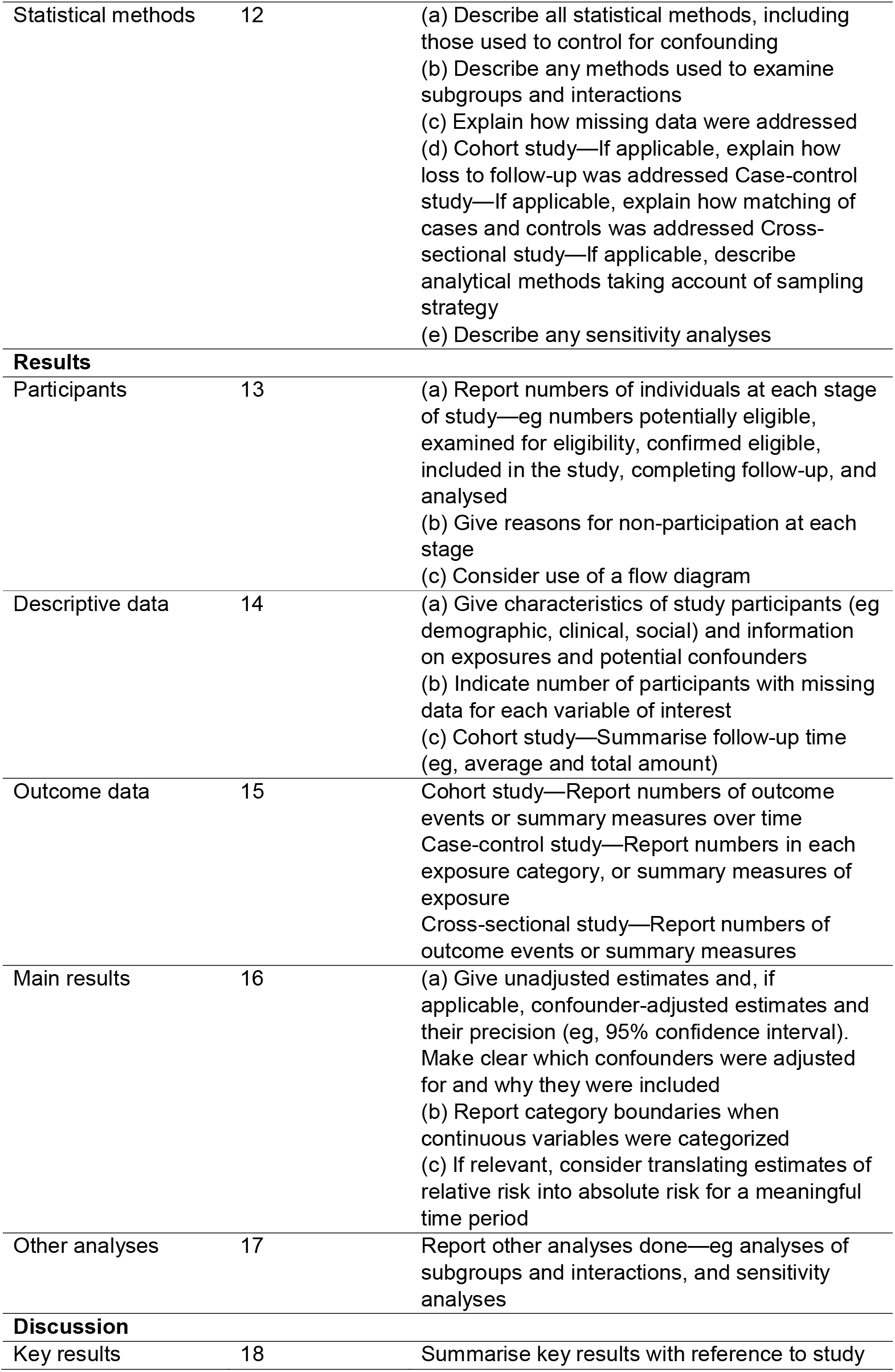
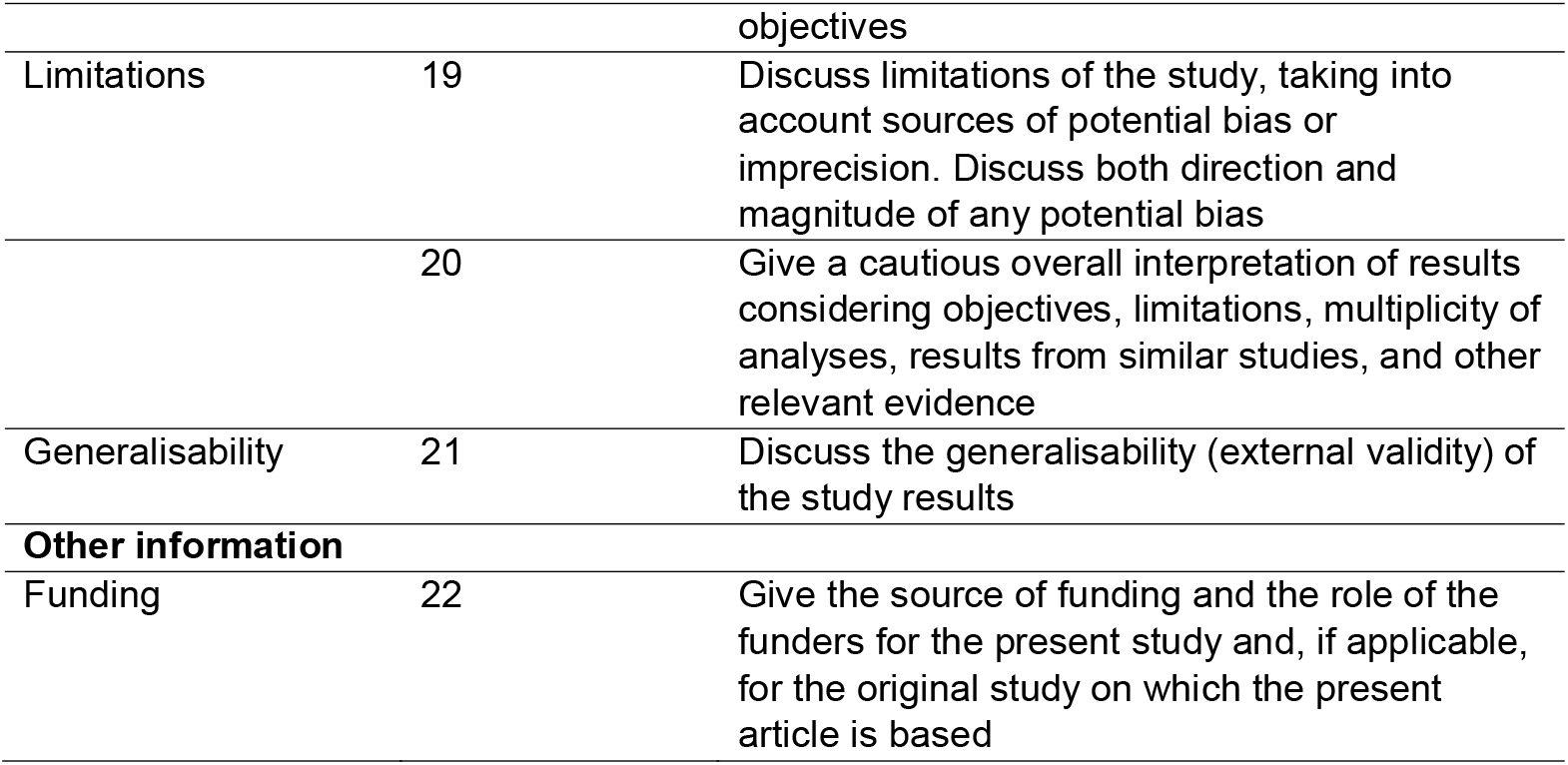

**SUPPORTING INFORMATION APPENDIX B** **STROBE Checklist of items that should be included in reports of observational studies**
|  | **Item** | **Recommendation** |
| --- | --- | --- |
| **Title and abstract** | 1 | (a) Indicate the study's design with a commonly used term in the title or the abstract<br>(b) Provide in the abstract an informative and balanced summary of what was done and what was found |
| **Introduction** |  |  |
| Background/rationale | 2 | Explain the scientific background and rationale for the investigation being reported |
| Objectives | 3 | State specific objectives, including any prespecified hypotheses |
| **Methods** |  |  |
| Study design | 4 | Present key elements of study design early in the paper |
| Setting | 5 | Describe the setting, locations, and relevant dates, including periods of recruitment, exposure, follow-up, and data collection |
| Participants | 6 | (a) Cohort study—Give the eligibility criteria, and the sources and methods of selection of participants. Describe methods of follow-up<br>Case-control study—Give the eligibility criteria, and the sources and methods of case ascertainment and control selection. Give the rationale for the choice of cases and controls<br>Cross-sectional study—Give the eligibility criteria, and the sources and methods of selection of participants<br>(b) Cohort study—For matched studies, give matching criteria and number of exposed and unexposed<br>Case-control study—For matched studies, give matching criteria and the number of controls per case |
| Variables | 7 | Clearly define all outcomes, exposures, predictors, potential confounders, and effect modifiers. Give diagnostic criteria, if applicable |
| Data sources/measurement | 8 | For each variable of interest, give sources of data and details of methods of assessment (measurement). Describe comparability of assessment methods if there is more than one group |
| Bias | 9 | Describe any efforts to address potential sources of bias |
| Study size | 10 | Explain how the study size was arrived at |
| Quantitative variables | 11 | Explain how quantitative variables were handled in the analyses. If applicable, describe which groupings were chosen and why |
| Statistical methods | 12 | (a) Describe all statistical methods, including those used to control for confounding<br>(b) Describe any methods used to examine subgroups and interactions<br>(c) Explain how missing data were addressed<br>(d) Cohort study—If applicable, explain how loss to follow-up was addressed Case-control study—If applicable, explain how matching of cases and controls was addressed Cross-sectional study—If applicable, describe analytical methods taking account of sampling strategy<br>(e) Describe any sensitivity analyses |
| **Results** |  |  |
| Participants | 13 | (a) Report numbers of individuals at each stage of study—eg numbers potentially eligible, examined for eligibility, confirmed eligible, included in the study, completing follow-up, and analysed<br>(b) Give reasons for non-participation at each stage<br>(c) Consider use of a flow diagram |
| Descriptive data | 14 | (a) Give characteristics of study participants (eg demographic, clinical, social) and information on exposures and potential confounders<br>(b) Indicate number of participants with missing data for each variable of interest<br>(c) Cohort study—Summarise follow-up time (eg, average and total amount) |
| Outcome data | 15 | Cohort study—Report numbers of outcome events or summary measures over time<br>Case-control study—Report numbers in each exposure category, or summary measures of exposure<br>Cross-sectional study—Report numbers of outcome events or summary measures |
| Main results | 16 | (a) Give unadjusted estimates and, if applicable, confounder-adjusted estimates and their precision (eg, 95% confidence interval). Make clear which confounders were adjusted for and why they were included<br>(b) Report category boundaries when continuous variables were categorized<br>(c) If relevant, consider translating estimates of relative risk into absolute risk for a meaningful time period |
| Other analyses | 17 | Report other analyses done—eg analyses of subgroups and interactions, and sensitivity analyses |
| **Discussion** |  |  |
| Key results | 18 | Summarise key results with reference to study |
|  |  | objectives |
| Limitations | 19 | Discuss limitations of the study, taking into account sources of potential bias or imprecision. Discuss both direction and magnitude of any potential bias |
|  | 20 | Give a cautious overall interpretation of results considering objectives, limitations, multiplicity of analyses, results from similar studies, and other relevant evidence |
| Generalisability | 21 | Discuss the generalisability (external validity) of the study results |
| **Other information** |  |  |
| Funding | 22 | Give the source of funding and the role of the funders for the present study and, if applicable, for the original study on which the present article is based |

## SUPPORTING INFORMATION APPENDIX C

### FULL DETAILS OF STATISTICAL ANALYSIS BY RESEARCH QUESTION

#### Research Question 1: Is SEP associated with vaping?

The primary sample with multiple imputations was used to create a stabilised inverse probability weight for the exposure (E) using our set of exposure-outcome confounders (x). This was based on the following two logistic regression models: (1) SEP as outcome, with no predictors (i.e. estimating: P(E)), and (2) SEP as outcome, with all exposure-outcome confounders as predictors (i.e. estimating: P(E|x)). This was used to calculate the predicted probabilities of each respondents’ observed exposure level (P*(E) and P*(E|x), where P*=P if the exposure=1 and P*=1-P if the exposure=0). Then, the exposure weight was calculated as P*(E)/P*(E|x).

Next, we created an inverse probability weight for w8 smoking status (M), conditioned on both exposure-outcome confounders (x), and mediator outcome confounders (z), using the following two logistic regression models: (1) wave 8 smoking status as outcome, predicted only by SEP (i.e. estimating: P(M|E)), and (2) wave 8 smoking status as outcome, predicted by SEP, and all exposure-outcome and mediator-outcome confounders (i.e. estimating: P(M|E, x, z)). This was conducted in two stages, with the first stage being conducted for ever vs. never smokers and the second stage for ever vs. current smokers. The vaping and smoking history variables were only included in the second stage as they were either highly correlated or deterministically-related to smoking status. This was used to calculate predicted probabilities (P*) of each respondents’ observed wave 8 smoking status, and the wave 8 smoking status weight is calculated as P*(M|E)/P*(M|E, x, z). This was multiplied together with the exposure weight (and the sampling/non-response weights) and represents respondents’ wave 9 analysis weight. Finally, a further weight was created for having smoking data at wave 10 (C), conditional on wave 9 smoking status (Y9). This was based on the following models: (1) Smoking data at wave 10 as outcome, predicted only by SEP, w8 smoking status and w9 smoking status (i.e. estimating: P(C|E, M, Y9)), and (2) Smoking data at wave 10 as outcome, predicted by SEP, w8 smoking status, w9 smoking status and all confounders (i.e. estimating: P(C|E, M, Y9, x, z)). An inverse probability weight was created using P* as above, and this is multiplied together with the wave 9 analysis weight to create a wave 10 analysis weight.

Logistic regression models were then used, with this final analysis weight applied, to examine the relationship between education and vaping, with wave 8 smoking status set to either current or ex-smoking. The odds ratio from these models can be interpreted as the CDE of education on vaping.

#### Research Question 2: Is vaping associated with smoking cessation?

In a similar process to research question 1, we created two inverse probability weights based on the following two logistic regression models: (1) wave 8 smoking status as outcome, predicted only by regular vaping status, and (2) wave 8 smoking status as outcome, predicted by regular vaping status and all exposure-outcome and mediator-outcome confounders. These weights were then multiplied together and used in logistic regression models, which estimate the CDE of regular vaping on smoking cessation/relapse.

#### Research Question 3: Is SEP associated with smoking cessation/relapse?

The same weights as research question 1 were used to estimate the CDE of education on smoking cessation/relapse.

#### Research Question 4: Does vaping mediate or moderate the associations between SEP and smoking cessation/relapse?

The same inverse probability weights used for research questions 1 and 2 were used, but with an additional step of weighting for regular vaping as the mediator. These weights were multiplied together and used in logistic regression models with smoking cessation/relapse as the outcome and SEP, wave 8 regular vaping, and the interaction between the two included as predictors. This estimates the CDE of education on smoking cessation/relapse with vaping status set to either regular vaping or not regular vaping.

## SUPPORTING INFORMATION APPENDIX D

Sensitivity analysis: vaping variable categorised as 0 = non-vaper, 1 = infrequent or regular vaper

**Table S2.** Estimates of Effects of SEP on Smoking Cessation/Relapse, With and Without Interaction by Vaping Status

|  | **Unadjusted Association**<br>**between Having No Degree**<br>**and Smoking**<br>**Cessation/Relapse** | **Controlled Direct Effect of**<br>**Having No Degree on**<br>**Smoking Cessation/Relapse** | **Controlled Direct Effect of**<br>**Having No Degree on**<br>**Smoking Cessation/Relapse**<br>**Among Non-Vapers** | **Controlled Direct Effect of**<br>**Having No Degree on**<br>**Smoking Cessation/Relapse**<br>**Among Vapers** |
| --- | --- | --- | --- | --- |
|  | OR [95% CI] | OR [95% CI] | OR [95% CI] | OR [95% CI] |
| **Smoking Cessation**<br>[Reference: Has Degree]<br>No Degree | 0.62 [0.52-0.73] | 0.65 [0.54-0.77] | 0.58 [0.48-0.72] | 1.23 [0.70-2.14] |
| **Smoking Relapse**<br>[Reference: Has Degree]<br>No Degree | 1.28 [0.93-1.75] | 1.67 [1.26-2.23] | 1.52 [0.99-2.33] | 1.74 [0.85-3.55] |

Notes: OR = odds ratio and 95% CI = 95% confidence interval. Vaping is defined as using or sometimes using e-cigarettes. The unadjusted association uses UKHLS wave 8 sampling weights to account for survey design and non-response but does not adjust for any confounders. The controlled direct effect uses inverse probability weighted marginal structural modelling to additionally adjust for exposure-outcome confounders and mediator-outcome confounders.

**Table S3.** Estimates of Effects of SEP on Vaping Among Current Smokers and Ex-Smokers

|  | **Unadjusted Association**<br>**between Having No Degree and**<br>**Vaping** | **Controlled Direct Effect of**<br>**Having No Degree on Vaping** |
| --- | --- | --- |
|  | OR [95% CI] | OR [95% CI] |
| **Wave 8 Vaping (Ex-Smokers)**<br>[Reference: Has Degree]<br>No Degree | 1.27 [1.03, 1.58] | 1.67 [1.35, 2.06] |
| **Wave 8 Vaping (Current Smokers)**<br>[Reference: Has Degree]<br>No Degree | 1.05 [0.82, 1.35] | 1.03 [0.79, 1.37] |

Notes: OR = odds ratio and 95% CI = 95% confidence interval. Vaping is defined as using or sometimes using e-cigarettes. The unadjusted association uses UKHLS wave 8 sampling weights to account for survey design and non-response but does not adjust for any confounders. The controlled direct effect uses inverse probability weighted marginal structural modelling to additionally adjust for exposure-outcome confounders and mediator-outcome confounders.

**Table S4.** Estimates of Effects of Vaping on Smoking Cessation/Relapse

|  | **Unadjusted Association**<br>**between Wave 8 Vaping and**<br>**Smoking Cessation/Relapse** | **Controlled Direct Effect of**<br>**Wave 8 Vaping on Smoking**<br>**Cessation/Relapse** |
| --- | --- | --- |
|  | OR [95% CI] | OR [95% CI] |
| **Smoking Cessation**<br>[Reference: Not Vaper]<br>Vaper | 1.12 [0.94-1.35] | 0.98 [0.76-1.27] |
| **Smoking Relapse**<br>[Reference: Not Vaper]<br>Vaper | 3.10 [2.32-4.13] | 3.27 [2.37-4.50] |

Notes: OR = odds ratio and 95% CI = 95% confidence interval. Vaping is defined as using or sometimes using e-cigarettes. The unadjusted association uses UKHLS wave 8 sampling weights to account for survey design and non-response but does not adjust for any confounders. The controlled direct effect uses inverse probability weighted marginal structural modelling to additionally adjust for exposure-outcome confounders and mediator-outcome confounders.

## SUPPORTING INFORMATION APPENDIX E

Sensitivity analysis: SEP based on NSSEC, categorised as 0 = management/professional, 1 = not management/professional

**Table S5.** Estimates of Effects of SEP on Smoking Cessation/Relapse, With and Without Interaction by Vaping Status

|  | Unadjusted Association<br>between NSSEC and<br>Smoking Cessation/Relapse | Controlled Direct Effect of<br>NSSEC on Smoking<br>Cessation/Relapse | Controlled Direct Effect of<br>NSSEC on Smoking<br>Cessation/Relapse<br>Among Non-Regular Vapers | Controlled Direct Effect of<br>NSSEC on Smoking<br>Cessation/Relapse<br>Among Regular Vapers |
| --- | --- | --- | --- | --- |
|  | OR [95% CI] | OR [95% CI] | OR [95% CI] | OR [95% CI] |
| **Smoking Cessation**<br>[Reference: Management/Professional]<br>Not Management/Professional | 0.75 [0.63-0.89] | 0.95 [0.76-1.18] | 0.93 [0.71-1.23] | 2.02 [0.80-5.13] |
| **Smoking Relapse**<br>[Reference: Management/Professional]<br>Not Management/Professional | 0.95 [0.69-1.33] | 1.30 [0.95-1.82] | 1.32 [0.76-2.29] | 1.61 [0.63-4.07] |

Notes: OR = odds ratio and 95% CI = 95% confidence interval. Regular vaping is defined as vaping at least weekly. The unadjusted association uses UKHLS wave 8 sampling weights to account for survey design and non-response but does not adjust for any confounders. The controlled direct effect uses inverse probability weighted marginal structural modelling to additionally adjust for exposure-outcome confounders and mediator-outcome confounders.

**Table S6.** Estimates of Effects of SEP on Regular Vaping Among Current Smokers and Ex-Smokers

|  | **Unadjusted Association**<br>**between NSSEC and Vaping** | **Controlled Direct Effect of**<br>**NSSEC on Vaping** |
| --- | --- | --- |
|  | OR [95% CI] | OR [95% CI] |
| **Wave 8 Regular Vaping (Ex-Smokers)**<br>[Reference: Management/Professional]<br>Not Management/Professional | 0.83 [0.66-1.04] | 1.09 [0.85-1.39] |
| **Wave 8 Regular Vaping (Current Smokers)**<br>[Reference: Management/Professional]<br>Not Management/Professional | 1.09 [0.77-1.54] | 1.01 [0.65-1.57] |

*Notes:* OR = odds ratio and 95% CI = 95% confidence interval. Regular vaping is defined as vaping at least weekly. The unadjusted association uses UKHLS wave 8 sampling weights to account for survey design and non-response but does not adjust for any confounders. The controlled direct effect uses inverse probability weighted marginal structural modelling to additionally adjust for exposure-outcome confounders and mediator-outcome confounders.

## SUPPORTING INFORMATION APPENDIX F

Sensitivity analysis: SEP based on NSSEC, categorised as 0 = in paid employment, 1 = not in paid employment

**Table S7.** Estimates of Effects of SEP on Smoking Cessation/Relapse, With and Without Interaction by Vaping Status

|  | Unadjusted Association<br>between NSSEC and<br>Smoking Cessation/Relapse | Controlled Direct Effect of<br>NSSEC on Smoking<br>Cessation/Relapse | Controlled Direct Effect of<br>NSSEC on Smoking<br>Cessation/Relapse<br>Among Non-Regular Vapers | Controlled Direct Effect of<br>NSSEC on Smoking<br>Cessation/Relapse<br>Among Regular Vapers |
| --- | --- | --- | --- | --- |
|  | OR [95% CI] | OR [95% CI] | OR [95% CI] | OR [95% CI] |
| **Smoking Cessation**<br>[Reference: in Paid Employment]<br>Not in Paid Employment | 0.80 [0.69-0.93] | 0.86 [0.73-1.01] | 0.88 [0.72-1.08] | 1.62 [0.89-2.92] |
| **Smoking Relapse**<br>[Reference: in Paid Employment]<br>Not in Paid Employment | 0.59 [0.45-0.78] | 1.05 [0.79-1.39] | 1.10 [0.71-1.69] | 0.67 [0.25-1.77] |

Notes: OR = odds ratio and 95% CI = 95% confidence interval. Regular vaping is defined as vaping at least weekly. The unadjusted association uses UKHLS wave 8 sampling weights to account for survey design and non-response but does not adjust for any confounders. The controlled direct effect uses inverse probability weighted marginal structural modelling to additionally adjust for exposure-outcome confounders and mediator-outcome confounders.

**Table S8.** Estimates of Effects of SEP on Regular Vaping Among Current Smokers and Ex-Smokers

|  | Unadjusted Association<br>between NSSEC and Vaping | Controlled Direct Effect of<br>NSSEC on Vaping |
| --- | --- | --- |
|  | OR [95% CI] | OR [95% CI] |
| **Wave 8 Regular Vaping (Ex-Smokers)**<br>[Reference: in Paid Employment]<br>Not in Paid Employment | 0.46 [0.37-0.57] | 0.67 [0.53-0.86] |
| **Wave 8 Regular Vaping (Current Smokers)**<br>[Reference: in Paid Employment]<br>Not in Paid Employment | 0.87 [0.66-1.14] | 0.70 [0.51-0.96] |

*Notes:* OR = odds ratio and 95% CI = 95% confidence interval. Regular vaping is defined as vaping at least weekly. The unadjusted association uses UKHLS wave 8 sampling weights to account for survey design and non-response but does not adjust for any confounders. The controlled direct effect uses inverse probability weighted marginal structural modelling to additionally adjust for exposure-outcome confounders and mediator-outcome confounders.

## SUPPORTING INFORMATION APPENDIX G

Sensitivity analysis: no qualifications used instead of no degree as the educational attainment SEP measure)

**Table S9.** Estimates of Effects of SEP on Regular Vaping Among Current Smokers and Ex-Smokers

|  | **Unadjusted Association between Having**<br>**No Qualifications and Regular Vaping** | **Controlled Direct Effect of Having No**<br>**Qualifications on Regular Vaping** |
| --- | --- | --- |
|  | OR [95% CI] | OR [95% CI] |
| **Wave 8 Regular Vaping (Ex-Smokers)**<br>[Reference: Has Qualifications]<br>No Qualifications | 0.69 [0.54-0.87] | 0.86 [0.67-1.11] |
| **Wave 8 Regular Vaping (Current Smokers)**<br>[Reference: Has Qualifications]<br>No Qualifications | 1.17 [0.88-1.55] | 1.03 [0.74-1.40] |

Notes: OR = odds ratio and 95% CI = 95% confidence interval. Regular vaping is defined as vaping at least weekly. The unadjusted association uses UKHLS wave 8 sampling weights to account for survey design and non-response but does not adjust for any confounders. The controlled direct effect uses inverse probability weighted marginal structural modelling to additionally adjust for exposure-outcome confounders and mediator-outcome confounders.

**Table S10.**
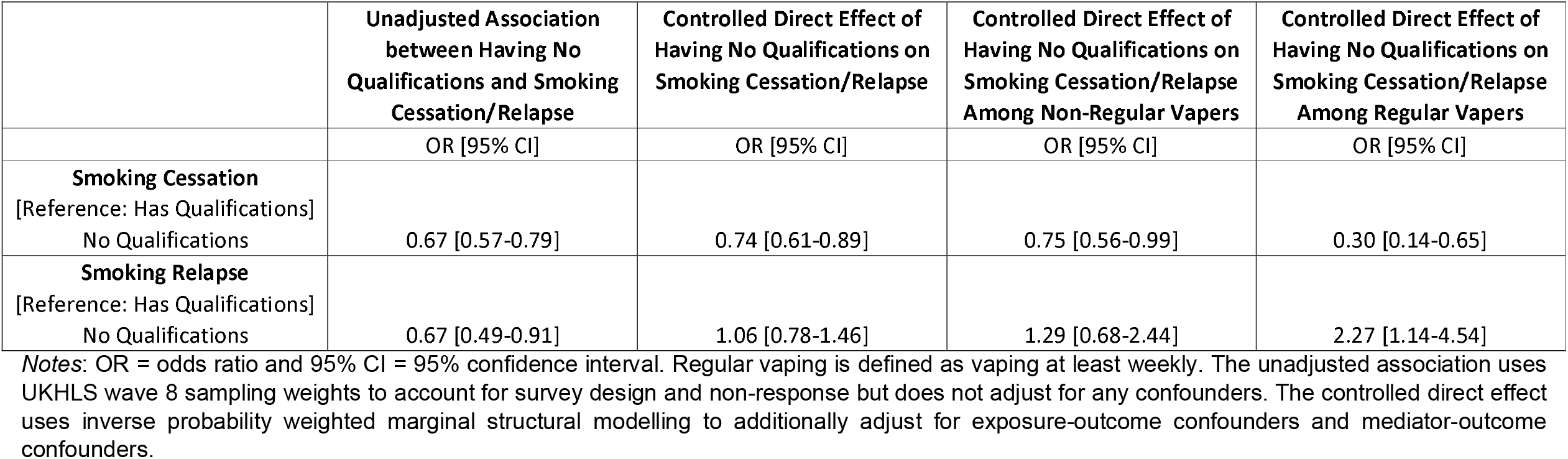
Estimates of Effects of SEP on Smoking Cessation/Relapse, With and Without Interaction by Regular Vaping Status

